# COVID-19 in the Mexican Social Security Institute (IMSS) population. Prevalent symptoms

**DOI:** 10.1101/2022.04.12.22273734

**Authors:** Arturo Juárez-Flores, Iván J. Ascencio-Montiel, Juan Pablo Gutiérrez, Stefano M. Bertozzi, Víctor H. Borja-Aburto, Gustavo Olaiz

## Abstract

**Background:** The disease caused by the new coronavirus SARS-CoV-2, COVID-19, which appeared in early 2020 in Mexico, was the second leading cause of mortality in the country that year and has generated an increasing demand for medical care. By January 2022, 4.13 million cases and 300 thousand direct deaths have been documented.

**Objective:** To describe the main symptoms of people with a positive test for SARS-CoV-2 treated at the Mexican Institute of Social Security (IMSS) by sex, age group, and IMSS delegation.

**Methods:** 4.5 million epidemiological reports were registered in the IMSS epidemiological surveillance system between February 2020 and November 2021. They were analyzed, reporting for those with either a positive PCR or rapid test, the prevalence of symptoms by sex, groups of age, and IMSS delegation.

**Results:** Among the population treated at the IMSS, six symptoms are observed as the most prevalent in general, as well as by sex, age groups, and state of residence: cefalea, fever, cough, myalgia, odynophagia, and arthralgias.

**Conclusions:** A better understanding of the clinical picture with which confirmed cases of COVID-19 present contributes to reporting timely diagnoses, considering the particularities by sex, age, and place of residence.

## Background

Severe acute respiratory syndrome coronavirus 2 (SARS-CoV-2) is an emerging pathogen that appeared in December 2019 in Wuhan, Hubei province, China (1, 2) that has generated a health and social crisis of major proportions with a pandemic that continues to be active to date.

By January 2022, 51.8 million cases have been confirmed globally, with about five million deaths. In Mexico, 4.13 million confirmed cases have been reported, and 300 thousand direct deaths have been registered (3).

In Mexico, a high lethality of COVID-19 has been documented, which has been related to the presence of co-morbidities and social inequality, aspects that have led to greater severity in cases (4, 5).

During the two years of evolution of the pandemic, COVID-19 has been described with diverse symptoms. It has been changing over time, probably related to the variants of the virus and the population’s profile.

COVID-19 presents peculiarities in the form of presentation since an extensive range of non-specific symptoms has been documented, and although predominantly respiratory, gastrointestinal symptoms are also relevant. This wide range of symptoms creates challenges producing a timely diagnosis in the absence of specific symptoms.

Various studies have documented the diversity of symptoms presented by people with COVID-19, with variations between countries; it has been proposed that there are groups of people with shared symptoms. In Mexico, a private COVID-19 testing provider study found three groups of patients: general symptoms, predominantly respiratory symptoms, and people among whom gastrointestinal symptoms were the central (6).

COVID-19 has become one of the leading health challenges in Mexico and has caused additional pressure on public health services that were already in challenging conditions. In this sense, it is relevant to have elements that contribute to planning care for people with this condition.

A better understanding of the symptoms will allow both to improve the opportunity for individual diagnosis, and better prepare the health services for the population’s profile. In this sense, analyzing the variations in symptomatology by sex, age, and place of residence can contribute to this objective.

This analysis focuses on the profile of symptoms reported by people suspected and confirmed to have COVID-19 treated at IMSS, Mexico’s leading social security institution and the country’s primary health services provider, identifying potential patterns by sex and age place of residence.

## Methods

This is a cross-sectional analysis of the data obtained through the IMSS Epidemiological Surveillance Online Notification System (SINOLAVE). To characterize the most overall symptomatology among the population in Mexico that the IMSS has attended, the distribution of symptoms is analyzed stratifying by sex, age, and place of residence.

### Data

The data for this analysis comes from the IMSS Epidemiological Surveillance Online Notification System (SINOLAVE), a registry platform of the leading health problems identified for epidemiological surveillance and that works in the 1,515 first-level medical units, 248 second-level hospitals, and ten national medical centers, and that is distributed in the 32 entities of the country. This report is updated at the same time as your clinical care and information on the initial assessment or medical consultation, hospital admission and stay data, tests for the detection of Sars-CoV-2 or other viral agents, discharge data, or death.

The registry of suspected cases of COVID-19 was incorporated into SINOLAVE from the beginning of the epidemic. In each unit, the person responsible for epidemiology validates and records the data from the epidemiological studies of suspected cases of COVID-19, which correspond to the initial assessment of suspected people, the results of detection tests, hospitalization data, and death of patients.

While most of the people treated in the IMSS units correspond to the population entitled to this social security institution, non-insured persons who, in the case of COVID-19, received care at the IMSS are also included.

The SINOLAVE comprises 50 variables and includes identification data (registration number for beneficiaries), sex, age if they speak an indigenous language, an outpatient or hospitalized case, the unit that diagnosis, and the care unit. It also includes the report of 12 comorbidities (obesity, diabetes, hypertension, COPD, asthma, immunosuppression, HIV, smoking, cardiovascular disease, tuberculosis, cancer, and kidney disease).

Likewise, it is recorded if the person had pneumonia and required intubation. On the other hand, it is recorded if the person received any antiviral and the date of death.

Regarding the signs and symptoms, the SINOLAVE captures 18 possible: fever, cough, headache, myalgia, odynophagia, arthralgia, rhinorrhea, malaise, chills, diarrhea, chest pain, anosmia, dysgeusia, abdominal pain, conjunctivitis, prostration, cyanosis, polypnea.

Among the advantages of SINOLAVE is the possibility of adapting to the various needs of the pandemic during its evolution and the presence of data validation before its integration.

The limitations of this data source correspond mainly to the learning curve in filling out the platform and the impossibility of discriminating between right holders and non-right holders.

### Analysis

The data was downloaded, stored, and cleaned in SQL server 2019, and the description of these was made in STATA 16.

The primary reported symptoms were identified considering a list of 18 possible ones captured in the SINOLAVE and the main comorbidities of a total of 12, both in general and by sex, age groups, and residence entity, reporting them as frequencies and percentages.

## Results

By November 20, 2021, 4.5 million people suspected of COVID-19 had been registered in SINOLAVE, of which 34% (1.53 million) were confirmed as cases either by PCR, rapid test, or epidemiological association (any death that had a positive contact but could not be verified by testing). Of those who tested positive, 49% were male, and their average age was 34 years. For all age groups, the percentage of women was higher compared to males (Table I).

**Table I.** Distribution of the population characteristics with confirmation of COVID-19 infection by test or epidemiologic association per sex

| Age Range | Man | Woman | Total |
| --- | --- | --- | --- |
| 00-05 Early infancy | 5,381 (47.0%) | 6,076 (53.0%) | 11,457 (100.0%) |
| 06-11 Infancy / School-age | 7,635 (48.1%) | 8,226 (51.9%) | 15,861 (100.0%) |
| 12-20 Adolescence | 40,292 (48.5%) | 42,862 (51.5%) | 83,154 (100.0%) |
| 21-30 Years | 186,772 (48.4%) | 199,089 (51.6%) | 385,861 (100.0%) |
| 31-40 Years | 178,925 (49.7%) | 181,093 (50.3%) | 360,018 (100.0%) |
| 41-50 Years | 144,414 (50.9%) | 139,457 (49.1%) | 283,871 (100.0%) |
| 51-60 Years | 91,456 (48.8%) | 95,985 (51.2%) | 187,441 (100.0%) |
| 61-70 Years | 52,857 (46.4%) | 61,060 (53.6%) | 113,917 (100.0%) |
| 71 Years or more | 39,925 (43.7%) | 51,540 (56.3%) | 91,465 (100.0%) |
| Total | 747,657 (100 %) | 785,388 (100 %) | 1,533,045 (100 %) |
| Frequency and percentage per sex, from total number of participants |  |  | p < 0.05 |

In supplementary Material 1, we report the comparison between the cases reported by the IMSS in the SINOLAVE database (red line) and the daily cases reported by the Mexican Ministry of Health (that includes those from SINOLAVE). As of November 2021, 3 waves of patients had been identified in Mexico, as shown in the figure.

Table II presents the data of the selected comorbidities. The most prevalent in both sexes are hypertension, obesity, and diabetes. Smoking is more frequent in men and asthma in women.

**Table II.** Distribution of comorbidities in COVID-19 confirmed cases per sex

| **Condition** | **Prevalence in women (%)**<br>**n=747,657** | **Prevalence in men (%)**<br>**n=785,388** | **Total (%)**<br>**n=1,533,045** |
| --- | --- | --- | --- |
| *Asthma* | 1.3% | 2.3% | 1.8% |
| *Diabetes* | 11.1% | 11.4% | 11.2% |
| *Cardiovascular Disease* | 1.3% | 1.0% | 1.2% |
| *EPOC* | 1.0% | 1.0% | 1.0% |
| *Hypertension* | 14.8% | 15.4% | 15.1% |
| *Immunosuppression* | 0.7% | 0.8% | 0.7% |
| *Obesity* | 11.3% | 13.2% | 12.2% |
| *Smoking* | 8.0% | 3.6% | 5.9% |
| *HIV* | 0.5% | 0.1% | 0.3% |
| *Prevalence by sex of the most common conditions* |  |  |  |

The states that have reported the largest number of cases attended by the IMSS are Mexico City and the State of Mexico, with almost a quarter of the reports. In contrast, the first five states in cases by 100 thousand inhabitants have been Baja California Sur (331.9), Colima (284.6), Mexico City (225.5), Quintana Roo (220.1), and Nayarit (210.3) (see Supplementary Material 3).

Regarding the symptoms reported at the time of registration in SINOLAVE, Table III reports the prevalence of the different symptoms among people with COVID-19. As can be seen, the symptom most frequently reported is headache (77.4% of cases), followed by cough (75.1%) and fever (64.7%). Other symptoms reported by more than half of those positive to SARS-CoV-2 are myalgia (60%), odynophagia (54.1%), and arthralgia (53.5%). Complementing the list of the ten most frequent are rhinorrhea (40.4%), attack to the general state (40.3%), shivers (34.7%), and anosmia (22%).

**Table III.** Prevalent symptomatology of confirmed COVID-19 cases by sex

| Condition | Prevalence in women (%)<br>n=747,657 | Prevalence in men (%)<br>n=785,388 | Total (%)<br>n=1,533,045 |
| --- | --- | --- | --- |
| ANOSMIA | 20.5% | 23.6% | 22.0% |
| ARTHRALGIA | 53.4% | 53.6% | 53.5% |
| GENERAL DEBILITATION | 41.2% | 39.4% | 40.3% |
| HEADACHE | 75.8% | 79.0% | 77.4% |
| CYANOSIS | 2.1% | 1.6% | 1.9% |
| CONJUNCTIVITIS | 7.2% | 7.8% | 7.5% |
| DIARRHEA | 14.2% | 13.8% | 14.0% |
| DYSGEUSIA | 19.1% | 21.8% | 20.4% |
| ABDOMINAL PAIN | 8.9% | 9.6% | 9.2% |
| THORACIC PAIN | 20.7% | 21.2% | 20.9% |
| SHIVERING | 34.7% | 34.8% | 34.7% |
| FEVER | 68.3% | 60.9% | 64.7% |
| MYALGIA | 59.9% | 60.1% | 60.0% |
| ODYNOPHAGIA | 53.0% | 55.4% | 54.1% |
| OTHERS | 2.2% | 2.3% | 2.3% |
| POLYPNEA | 2.1% | 1.6% | 1.9% |
| RECUMBENCY | 4.6% | 4.1% | 4.4% |
| RINORRHEA | 38.4% | 42.5% | 40.4% |
| COUGH | 75.3% | 74.9% | 75.1% |
| **Prevalence and 95% confidence intervals by sex of the most common conditions** |  |  |  |

When comparing age groups, differences are observed in the relative order and prevalence of symptoms (Table IV), although the list for each group includes the same symptoms. Among children between 5 and 11 years of age, the five main symptoms are fever (76%), cough (63.5%), rhinorrhea (50.3%), headache (39.1%), and odynophagia (33.6). For adolescents aged 12 to 19 years, the five main symptoms are headache (71.6%), fever (69.2%), cough (67.2%), odynophagia (53.4%), rhinorrhea (48%), and myalgia (41.6%).

**Table IV.** Prevalent symptomatology per age group

| Symptom | Infants<br>(5 to 11<br>years of<br>age) | Adolescen<br>ts (12 to<br>19 years<br>of age) | Adults<br>20 to 30 | 31 to 50<br>years of<br>age | 51 years<br>of age and<br>older |
| --- | --- | --- | --- | --- | --- |
| ANOSMIA | 3.1% | 8.5% | 23.4% | 26.7% | 24.0% |
| ARTHRALGIA | 18.8% | 34.4% | 46.2% | 51.5% | 55.8% |
| GENERAL DEBILITATION | 30.1% | 29.6% | 31.7% | 34.8% | 39.0% |
| HEADACHE | 39.1% | 71.6% | 80.1% | 80.5% | 79.3% |
| CYANOSIS | 1.8% | 0.5% | 0.5% | 0.7% | 1.3% |
| CONJUNCTIVITIS | 7.5% | 8.0% | 7.7% | 8.1% | 7.9% |
| DIARRHEA | 14.9% | 9.6% | 11.0% | 13.7% | 14.0% |
| DYSGEUSIA | 3.0% | 7.9% | 21.2% | 24.5% | 22.2% |
| ABDOMINAL PAIN | 9.3% | 11.1% | 8.0% | 8.2% | 8.8% |
| THORACIC PAIN | 4.9% | 6.6% | 11.9% | 15.3% | 20.4% |
| SHIVERING | 16.3% | 24.8% | 30.2% | 33.6% | 36.1% |
| FEVER | 76.0% | 69.2% | 63.8% | 63.8% | 64.3% |
| MYALGIA | 23.1% | 41.6% | 54.3% | 59.3% | 62.4% |
| ODYNOPHAGIA | 33.6% | 53.4% | 59.2% | 58.5% | 56.2% |
| OTHERS | 2.3% | 1.6% | 1.7% | 1.9% | 2.2% |
| POLYPNEA | 1.8% | 0.5% | 0.5% | 0.7% | 1.3% |
| RECUMBENCY | 2.6% | 2.4% | 2.6% | 2.7% | 3.8% |
| RHINORRHEA | 50.3% | 48.0% | 47.7% | 45.9% | 41.6% |
| COUGH | 63.5% | 67.2% | 70.9% | 72.9% | 75.5% |

For people aged 20 to 30 years, the five main symptoms were headache (80.1%), cough (70.9%), fever (63.8%), odynophagia (59.2%), myalgia (54.3%), and rhinorrhea (47.7%). Among those between 31 and 50 years of age, the five most prevalent symptoms were headache (80.5%), cough (72.9%), fever (63.8%), myalgia (59.3%), and sore throat (58.5%).

Finally, for those aged 51 and over, the five most common symptoms were headache (79.3%), cough (75.5%), fever (64.3%), myalgia (62.4%), sore throat (56.2%), and arthralgia (55.8%).

Table V shows the five symptoms with the highest prevalence in each state. As can be seen, in 29 of the 32 states, the most prevalent symptom was headaches, and in 2 of the three states in which it was not the most prevalent, it ranked second. Among states, headaches ranged from 65.2% in Guerrero to 86.9% in Morelos.

**Table V.** Prevalent symptomatology per delegation

| State | 1 | 2 | 3 | 4 | 5 |
| --- | --- | --- | --- | --- | --- |
| Aguascalientes | Headache (73.9 %) | Cough (66.6 %) | Myalgia (55.1 %) | Odynophagia (53.6 %) | Fever (51.2 %) |
| Baja California | Headache (72.8 %) | Cough (71.2 %) | Fever (57.7 %) | Myalgia (57.6 %) | Arthralgia (49.9 %) |
| Baja California Sur | Headache (85.6 %) | Cough (79.9 %) | Fever (73.5 %) | Odynophagia (64.8 %) | Myalgia (61.3 %) |
| Campeche | Fever (74.5 %) | Cough (74.5 %) | Headache (71.8 %) | Odynophagia (52.5 %) | Myalgia (51.2 %) |
| Chiapas | Cough (83.5 %) | Headache (82.9 %) | Fever (78.6 %) | Myalgia (75.4 %) | Arthralgia (68.3 %) |
| Chihuahua | Headache (72.7 %) | Cough (71.8 %) | Myalgia (63.7 %) | Arthralgia (58.3 %) | Fever (58.0 %) |
| Ciudad de México | Headache (75.7 %) | Cough (75.5 %) | Fever (59.6 %) | Myalgia (58.3 %) | Odynophagia (54.8 %) |
| Coahuila | Headache (74.3 %) | Cough (70.2 %) | Fever (57.6 %) | Myalgia (52.7 %) | Arthralgia (47.2 %) |
| Colima | Headache (82.2 %) | Cough (77.6 %) | Fever (71.7 %) | Myalgia (64.2 %) | Arthralgia (59.1 %) |
| Durango | Headache (73.6 %) | Cough (65.8 %) | Fever (55.8 %) | Myalgia (48.5 %) | Arthralgia (44.3 %) |
| Estado de México | Headache (76.7 %) | Cough (76.3 %) | Fever (62.0 %) | Myalgia (59.4 %) | Odynophagia (54.2 %) |
| Guanajuato | Headache (84.2 %) | Cough (80.4 %) | Myalgia (66.5 %) | Fever (63.9 %) | Odynophagia (62.1 %) |
| Guerrero | Headache (65.2 %) | Cough (63.5 %) | Fever (62.9 %) | Myalgia (56.2 %) | Arthralgia (51.4 %) |
| Hidalgo | Headache (83.4 %) | Cough (82.3 %) | Fever (64.7 %) | Myalgia (62.7 %) | Odynophagia (59.4 %) |
| Jalisco | Headache (78.9 %) | Cough (78.7 %) | Fever (66.8 %) | Myalgia (63.8 %) | Arthralgia (58.9 %) |
| Michoacán | Headache (83.9 %) | Cough (81.9 %) | Fever (73.5 %) | Myalgia (64.0 %) | Arthralgia (61.7 %) |
| Morelos | Headache (86.9 %) | Cough (86.3 %) | Fever (78.7 %) | Myalgia (76.8 %) | Arthralgia (71.4 %) |
| Nayarit | Headache (82.2 %) | Cough (76.7 %) | Fever (70.3 %) | Myalgia (63.1 %) | Arthralgia (58.6 %) |
| Nuevo León | Headache (74.7 %) | Cough (69.2 %) | Fever (61.8 %) | Myalgia (54.6 %) | Arthralgia (47.1 %) |
| Oaxaca | Headache (81.6 %) | Cough (77.8 %) | Fever (74.5 %) | Myalgia (65.7 %) | Odynophagia (61.4 %) |
| Puebla | Cough (76.0 %) | Headache (75.4 %) | Fever (64.1 %) | Myalgia (58.9 %) | Odynophagia (57.9 %) |
| Querétaro | Headache (78.0 %) | Cough (74.4 %) | Odynophagia (60.7 %) | Myalgia (56.6 %) | Fever (55.5 %) |
| Quintana Roo | Headache (78.1 %) | Fever (75.6 %) | Cough (73.8 %) | Odynophagia (59.9 %) | Myalgia (51.9 %) |
| San Luis Potosí | Headache (75.7 %) | Cough (75.6 %) | Fever (60.8 %) | Myalgia (59.5 %) | Arthralgia (53.2 %) |
| Sinaloa | Headache (79.2 %) | Cough (74.4 %) | Fever (72.9 %) | Myalgia (65.8 %) | Arthralgia (62.2 %) |
| Sonora | Headache (79.0 %) | Cough (74.4 %) | Fever (69.4 %) | Myalgia (67.7 %) | Arthralgia (64.4 %) |
| Tabasco | Headache (83.1 %) | Cough (78.1 %) | Fever (77.3 %) | Myalgia (59.0 %) | Arthralgia (55.0 %) |
| Tamaulipas | Headache (75.4 %) | Cough (73.9 %) | Fever (69.2 %) | Myalgia (59.9 %) | Arthralgia (54.9 %) |
| Tlaxcala | Headache (80.9 %) | Cough (76.8 %) | Odynophagia (68.9 %) | Fever (59.4 %) | Myalgia (58.9 %) |
| Veracruz | Headache (78.9 %) | Cough (77.1 %) | Fever (74.1 %) | Myalgia (64.5 %) | Arthralgia (56.7 %) |
| Yucatán | Headache (73.9 %) | Cough (72.7 %) | Fever (71.2 %) | Myalgia (57.7 %) | Odynophagia (52.5 %) |
| Zacatecas | Headache (73.3 %) | Cough (68.7 %) | Myalgia (56.6 %) | Arthralgia (52.5 %) | Fever (50.2 %) |
| Total | HEADACHE (77.4%) | COUGH (75.1%) | FEVER (64.7%) | MYALGIA (60.0%) | ODYNOPHAGIA (54.1%) |

Similarly, the cough was the second most prevalent symptom in 29 of the 32 states, ranging between delegations from 63.5% in Guerrero to 83.5% in Chiapas. In this latter entity, the cough was the most prevalent symptom.

In 24 of the 32 states, the third most prevalent symptom has been fever, with a range between entities of 50.2% in Zacatecas –fever was the fifth most prevalent symptom— to 78.7% Morelos.

Myalgias, odynophagia, and arthralgias complete the list for all delegations regarding the five most prevalent symptoms. Six conditions in the 32 states are the main five symptoms among those individuals with COVID-19.

## Discussion

This analysis identified differences in the prevalence of symptoms by age. However, in general, in the different age groups of people treated at the IMSS with a diagnosis of COVID-19, the six main symptoms remain headache, cough, fever, myalgia, odynophagia, and arthralgia.

Likewise, these six symptoms make up the list of the five most prevalent among people diagnosed with COVID-19 in the 38 IMSS delegations, although with substantial differences in terms of their prevalence.

The pattern of symptoms identified for the Mexican population treated at the IMSS is consistent with those reported in other studies, although with differences that are relevant to highlight. A systematic review of studies published between January and April 2020 found that the five most frequently reported symptoms are fever (81.2%), cough (58.5%), fatigue (38.5%), dyspnea (26.1%), and sputum (25.8%) (7); that is, while fever and cough are also among the top five in Mexico, the high prevalence of headache stands out, which is not reported in the global review.

In an analysis of moderate cases of COVID-19 in Wuhan, it was documented that the main symptoms observed were fever, cough, and dyspnea (8); therefore, the high prevalence of headaches also stands out for the Mexican case. In one of the first published descriptive studies on patients from Hubei, China, the following prevalence of accompanying symptoms were described: loss of appetite (78.6%), diarrhea (34%), vomiting (3.9%), and abdominal pain (1.9%) (9).

In a study conducted in the United States, the prevalence of gastrointestinal symptoms in patients with COVID-19 is summarized from different studies, identifying the presence of anorexia between 12.2 and 50.2% of cases (10). An adjusted prevalence of 7.8% in outpatients and 22.7% in hospitalized patients is estimated regarding nausea and vomiting. (10).

In a review of the clinical presentation of COVID-19 in Africa, it was documented that the most prevalent symptoms, similar to the Mexican case, were fever (42.8%), cough (33.3%), headache (11.3%), and respiratory problems (16.8%)(11).

Regarding diarrhea, a prevalence of 5 to 10% of cases in outpatients and up to 36.6% of hospitalized patients in a region of Hubei, China is estimated. The prevalence of abdominal pain was also 3.6 to 6.8% of cases. Ageusia or dysgeusia had a high prevalence, with a value of 49.8% of cases.

Age influences the observed prevalence of severe symptoms such as prostration and dyspnea, while non-serious symptoms (headache, anosmia) are more frequent in the young population.

The proportions of comorbidities in patients differ by sex, with higher diabetes, asthma, and obesity in women and COPD, smoking, and cardiovascular disease in men.

The strengths of this study lie in its high number of observations and national representativeness since 30.1% of the cases recognized in the national database come from this institution; however, there are limitations to these data, such as the differences in the care and registration process in each of the medical units.

Subsequent evaluations will confirm the prevalent symptomatology and characterize the patient suspected of COVID-19.

## Conclusions

COVID 19 is an emerging disease that has become the second cause of death in Mexico during 2020 and one of the main reasons for health care. Considering the magnitude of the challenge it represents and the dynamics that have been documented, a permanent study of its symptoms is necessary to keep health professionals updated on the profile of the cases, both for their identification and for their management.

Subsequent works will focus on the evolution of symptoms in the population by the pandemic period, their relationship with diagnostic tests, and the outcome of cases.

## Data Availability

All data produced in the present study are available upon reasonable request to the auth

## Declaration of the absence of conflict of interest

Authors declare not having conflicts of interest.

## Supplementary Materials

**Supplementary Material 1.**
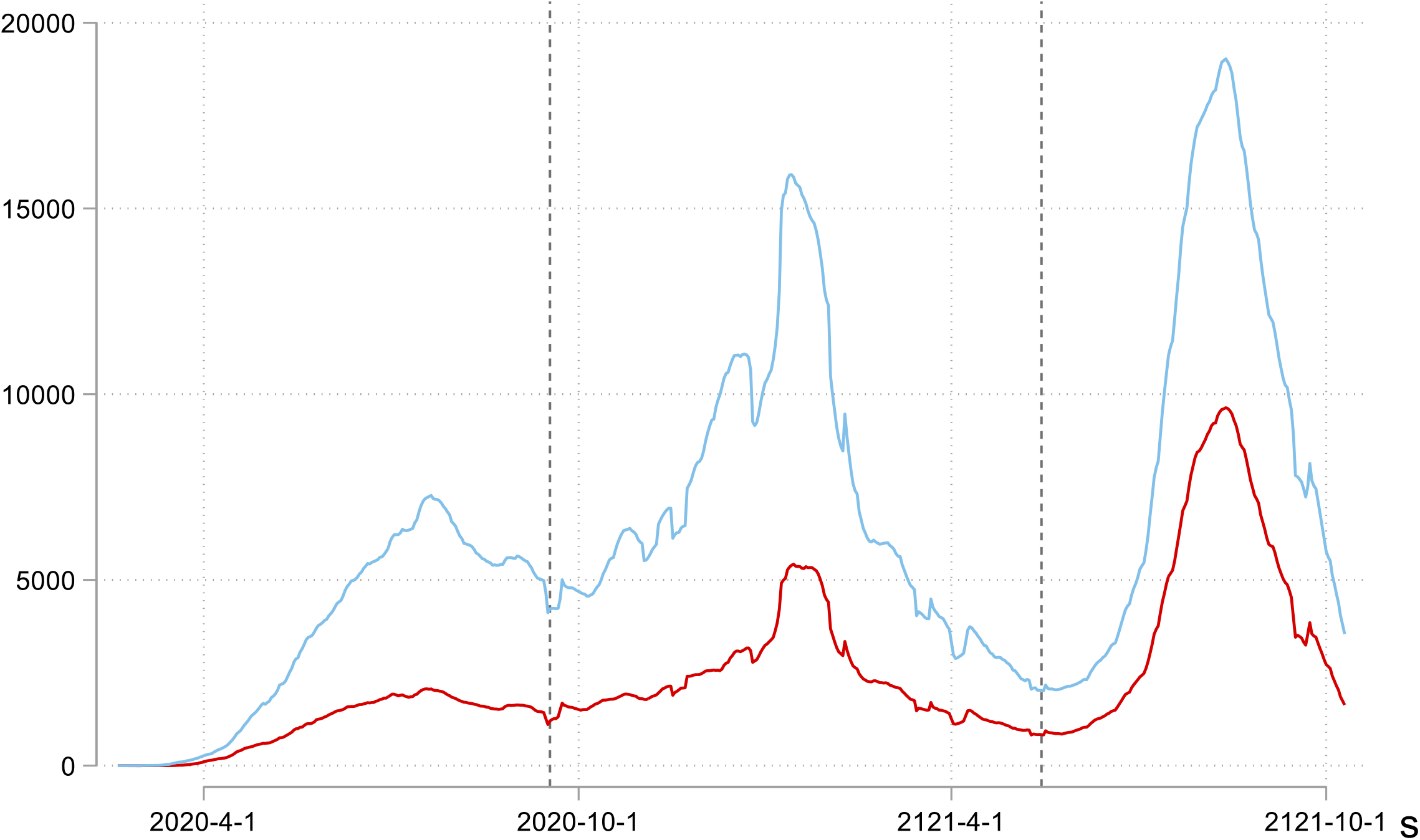
Confirmed COVID-19 cases by date of symptom onset treated by the IMSS. Moving average of 7 days. The red line represents the number of cases treated by the IMSS and the blue line the number of national cases.

**Supplementary Material 2.**
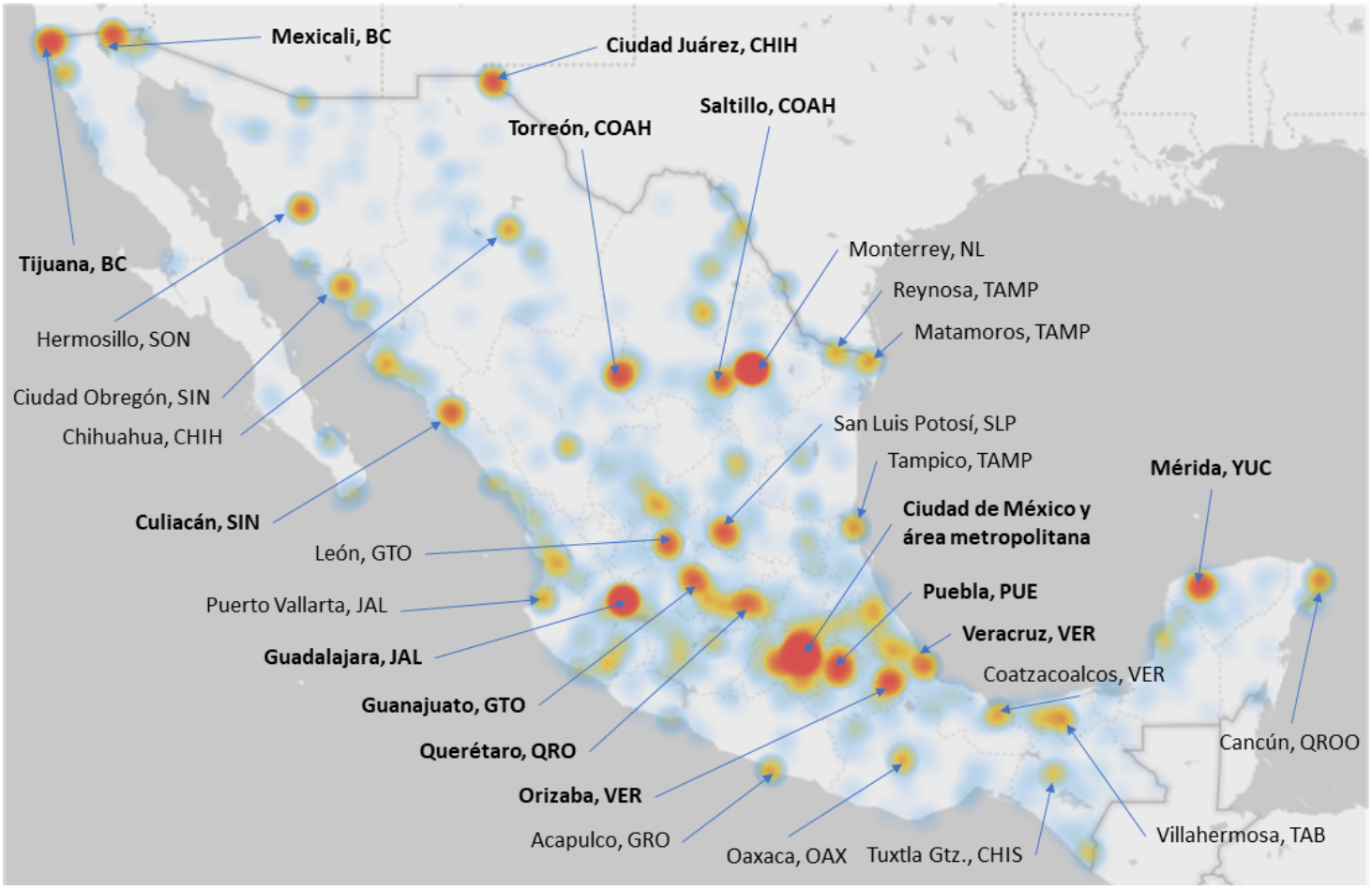
Density map with the distribution of confirmed COVID-19 cases. The regions in red represent medical units with maximum observed incidence, the regions in yellow units with 50% maximum and the regions in blue 25%

**Supplementary Material 3.**
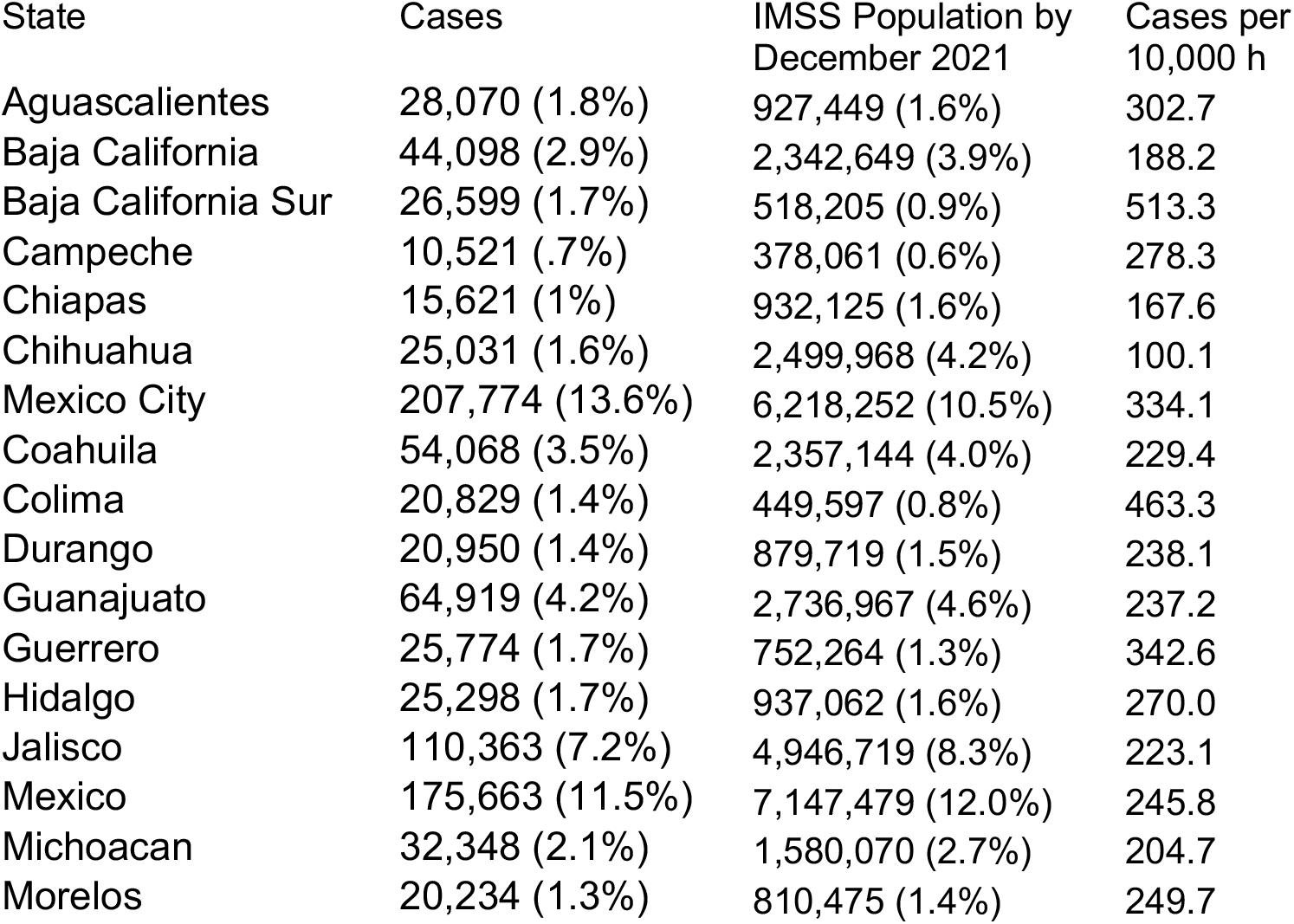
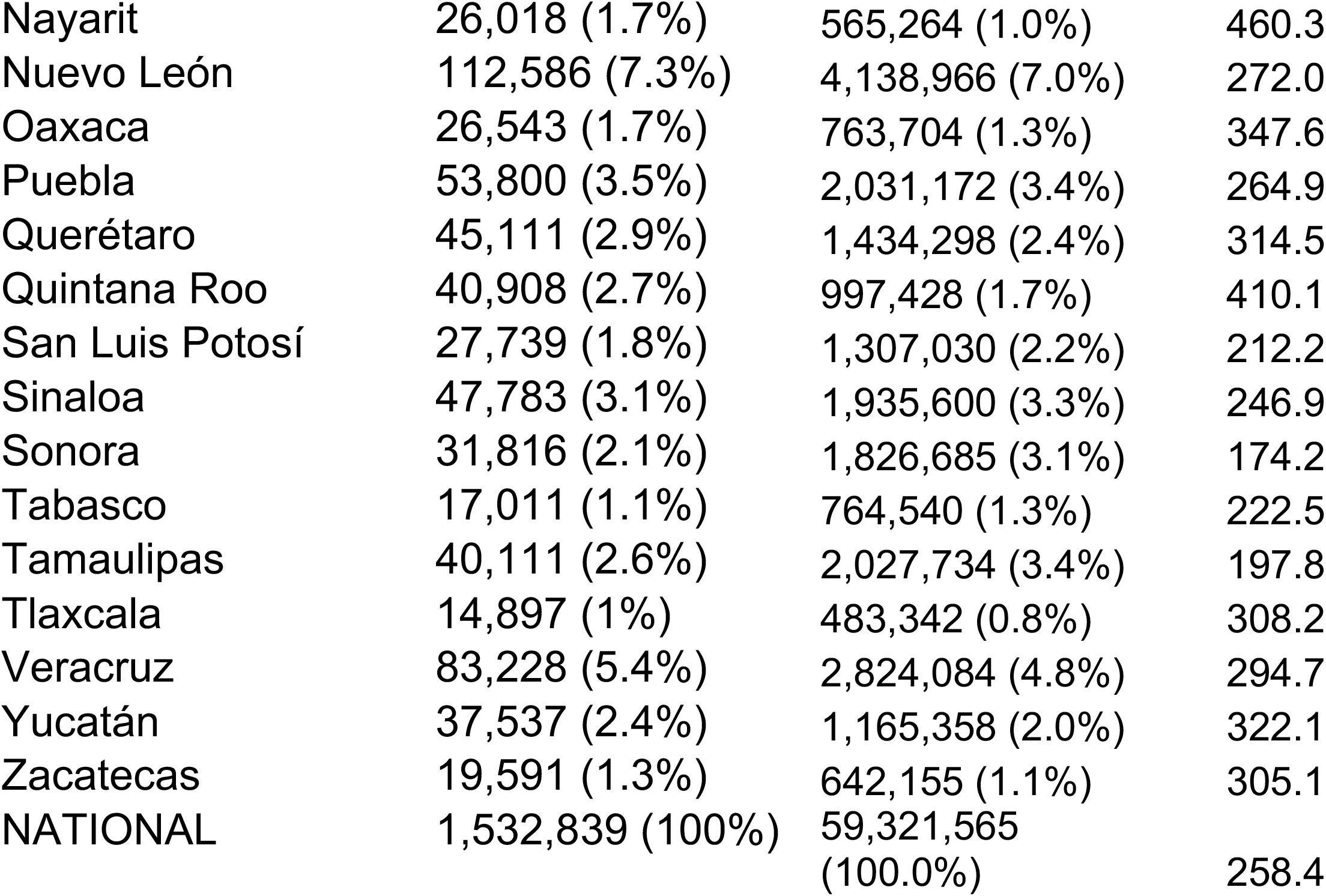
Percentage of confirmed cases per entity

